# Genomic monitoring unveils a high prevalence of SARS-CoV-2 Omicron variant in vaccine breakthrough cases

**DOI:** 10.1101/2022.02.16.22271059

**Authors:** Gúbio Soares Campos, Marta Giovanetti, Laise de Moraes, Helena Souza da Hora, Keila Oliveira Motta de Alcantara, Silvia Ines Sardi

## Abstract

Genome sequencing proved to be an excellent tool to monitor the molecular epidemiology of the disease caused by SARS-CoV-2, i.e., coronavirus disease (COVID-19). Some reports of infected, vaccinated individuals have aroused great interest because they are primarily being infected with circulating variants of concern (VOCs). To investigate the cases of infected, vaccinated individuals in Salvador, Bahia, Brazil, we performed genomic monitoring to estimate the magnitude of the different VOCs in these cases. Nasopharyngeal swabs from infected (symptomatic and asymptomatic), fully vaccinated individuals (n=29) who were of varying age and had RT-qPCR Ct values of ≤30 were subjected to viral sequencing using Nanopore technology. Our analysis revealed that the Omicron variant was found in 99% of cases and that only one case was due to the Delta variant. Infected, fully vaccinated patients have a favorable clinical prognosis; however, within the community, they become viral carriers with the aggravating factor of viral dissemination of VOCs not neutralized by the vaccines.

## Introduction

Severe acute respiratory syndrome coronavirus 2 (SARS-CoV-2), which causes COVID-19, a disease that emerged in December 2019 in Wuhan, China, fueled worldwide efforts to develop vaccines to control the rapid spread of infection.

Currently, there are several vaccines against SARS-CoV-2 approved by the World Health Organization (WHO)^1^. However, the viral replication of this RNA virus has general characteristics common to other RNA viruses; the high mutation rate allows it to generate new viral variants to improve its chances of survival in the host and escape to immune detection^2^.

It has been shown that SARS-CoV-2 vaccines do not protect against viral infection, although they significantly reduce morbidity and mortality in infected patients. Moreover, the neutralizing antibodies persist for no more than 4 or 5 months, even with a full two-dose regimen^3^. These factors may contribute to the fact that fully vaccinated individuals, as well as those with booster doses, may acquire the infection. Although the clinical situation of the infected patient who is fully vaccinated has a very favorable prognosis, they become a possible viral carrier and can trigger viral dissemination within the community^4,5^.

Recent studies of infected, vaccinated individuals have generated notable interest because they show that these individuals can primarily be infected with the circulating variants of concern (VOCs), such as Omicron (B.1.529). This variant is more aggressive, with greater transmissibility and infectivity compared to the Delta variant^6^.

Here, we conducted a genomic study to estimate the magnitude and range of SARS-CoV-2 VOCs in cases of infected (symptomatic and asymptomatic), fully vaccinated individuals.

## Materials and Methods

### Molecular detection of SARS-CoV2

Patients (n=29) of any gender or age group, symptomatic or not, after consenting, clinical information and oropharyngeal and nasal swab were collected at the Laboratory of Virology at Federal University of Bahia. Detection of SARS-CoV-2 was carried out using an RT-qPCR (GoTaq® Probe 1-Step RT-qPCR System; Promega, USA) assay following the CDC 2019 Novel Coronavirus (2019-nCoV) Real-Time Reverse Transcriptase (RT)– PCR Diagnostic Panel^7^.

### Ethics statement

This research was reviewed and approved by the Ethical Committee of the Federal University of Bahia (CAAE 30687320.9.0000.5662), and informed consent of all participating subjects or their legal guardians have been obtained.

### cDNA synthesis and whole genome sequencing

Samples (n=29) were selected for sequencing based on the Ct value (≤30) and availability of epidemiological metadata (sex, age, residence in Salvador, symptoms, etc.) (**Table 1**). The preparation of SARS-CoV-2 genomic libraries was performed using the nanopore sequencing technology^8^. The SuperScript IV Reverse Transcriptase kit (Invitrogen, USA) was initially used for cDNA synthesis, following the manufacturer’s instructions. The cDNA generated was subjected to multiplex PCR sequencing using the Q5 High Fidelity Hot-Start DNA Polymerase (New England Biolabs, UK) and a set of specific primers designed by the ARTIC Network for sequencing the complete SARS-CoV-2 genome (version 4)^9^. All experiments were performed in a biosafety level-2 cabinet. Amplicons were purified using 1x AMPure XP Beads (Beckman Coulter, USA) and quantified on a Qubit 3.0 fluorimeter (ThermoFisher Scientific, USA) using Qubit™ dsDNA HS Assay Kit (ThermoFisher Scientific, USA). DNA library preparation was performed using the Ligation Sequencing Kit SQK-LSK109 (Oxford Nanopore Technologies, UK) and the Native Barcoding Kit (EXP-NBD104 and EXP-NBD114, Oxford Nanopore Technologies, UK). Sequencing libraries were loaded into an R9.4 flow cell (Oxford Nanopore Technologies, UK). In each sequencing run, we used negative controls to prevent and check for possible contamination with less than 2% mean coverage.

**Table 1.**
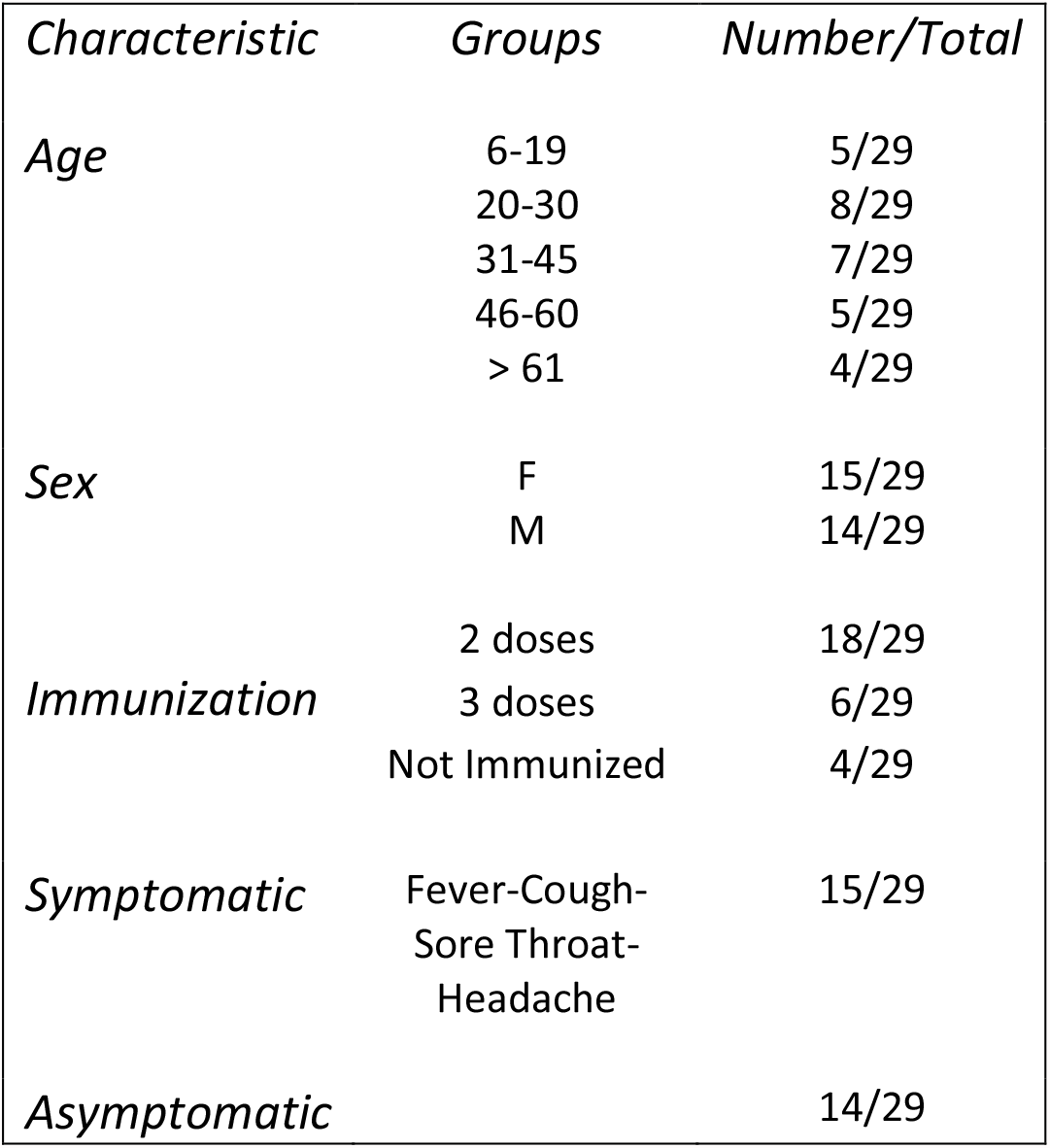
Demographic and Clinical characteristics in vaccinated or unvaccinated study group.

| *Characteristic* | *Groups* | *Number/Total* |
| --- | --- | --- |
| *Age* | 6-19 | 5/29 |
|  | 20-30 | 8/29 |
|  | 31-45 | 7/29 |
|  | 46-60 | 5/29 |
|  | > 61 | 4/29 |
| *Sex* | F | 15/29 |
|  | M | 14/29 |
| *Immunization* | 2 doses | 18/29 |
|  | 3 doses | 6/29 |
|  | Not Immunized | 4/29 |
| *Symptomatic* | Fever-Cough-Sore Throat-Headache | 15/29 |
| *Asymptomatic* |  | 14/29 |

### Generation of consensus sequences from nanopore

The fast5 files generated during sequencing were basecalled under the high-accuracy model performed by Guppy v.6.0.1 (Oxford Nanopore Technologies, UK). The basecalled fastQ files with a minimum Q score of 7 were selected for subsequent trim adaptor and demultiplex processes performed by Guppy v.6.0.1. Furthermore, the fastQ files were submitted to the ARTIC Network’s field bioinformatics pipeline v.1.2.1^9^. Briefly, a length filtering was performed to remove additional chimeras using artic guppyplex. The assembly was performed by Minimap2 v.2.17^10^ using Genbank accession no. MN908947.3 as genome reference. The primer sequences were trimmed with align_trim.py, the assembly was then polished, and the variant calling was performed by Medaka v.1.0.3 (Oxford Nanopore Technologies, UK) and evaluated by LongShot v.0.4.1^11^. The consensus sequences were then masked with “N” at regions with coverage depth <20, and the variant candidates were incorporated into the consensus genome using BCFtools v.1.10.2^12^. The alignment statistics were calculated with SAMtools v.1.10 (using htslib 1.10.2)^12^, exonerate v.2.4.0 (using glib version v.2.68.0)^13^ and Seqtk v.1.3-r106^14^. This entire workflow is available at https://github.com/khourious/vgapONT.

### Phylogenetic inference

Lineage assignment was performed using the Pangolin lineage classification software tool^15^. The newly identified isolates were compared to a diverse pool of genome sequences (n= 3,441) sampled worldwide collected up to October 28th, 2021. All sequences were aligned using the ViralMSA tool v.1.1.20^16,17^, and IQ-TREE2 v.2.2.0^18^ was used for phylogenetic analysis using the maximum likelihood approach. TreeTime v.0.8.5^19^ was used to transform this ML tree topology into a dated tree using a constant mean rate of 8.0 × 10^−4^ nucleotide substitutions per site per year, after the exclusion of outlier sequences.

## Results

In this study, all individuals (n=29) were vaccinated with a complete two-dose regimen, except for four 7-and 11-year-olds who were not vaccinated (Table 1). The symptoms self-referred by participants were mild or asymptomatic, without the need for hospital care. The RT-qPCR Ct values varied between 14.6 and 27.2, even in individuals who declared themselves as asymptomatic (Fig. 1). From those vaccinated individuals (n=25) 24 confirm the presence of Omicron variant and only one Delta variant. From unvaccinated individuals (n=4) confirmed an Omicron Variant’s infection. Our genomic surveillance analysis revealed that in the state of Bahia, three VOCs primarily dominated the epidemiology of the state. The Gamma variant (P.1) that was prominent during the second epidemic wave persisted until August 2021, when it was replaced by Delta (B.1.617.2) (Fig. 2a), which in turn was replaced by the emerging Omicron variant in December 2021. The Alpha variant was also detected in the state of Bahia at the end of December 2021, but it remained at a very low frequency (less than 1%). To explore the relationship between the sequenced genomes, we also constructed a phylogenetic tree with these VOCs and those from other parts of the world. Our time-stamped phylogeny revealed that the Omicron isolates in the Bahia state are scattered throughout the phylogeny, suggesting that multiple independent introductions have occurred through time (Fig. 2b).

**Figure 1.**
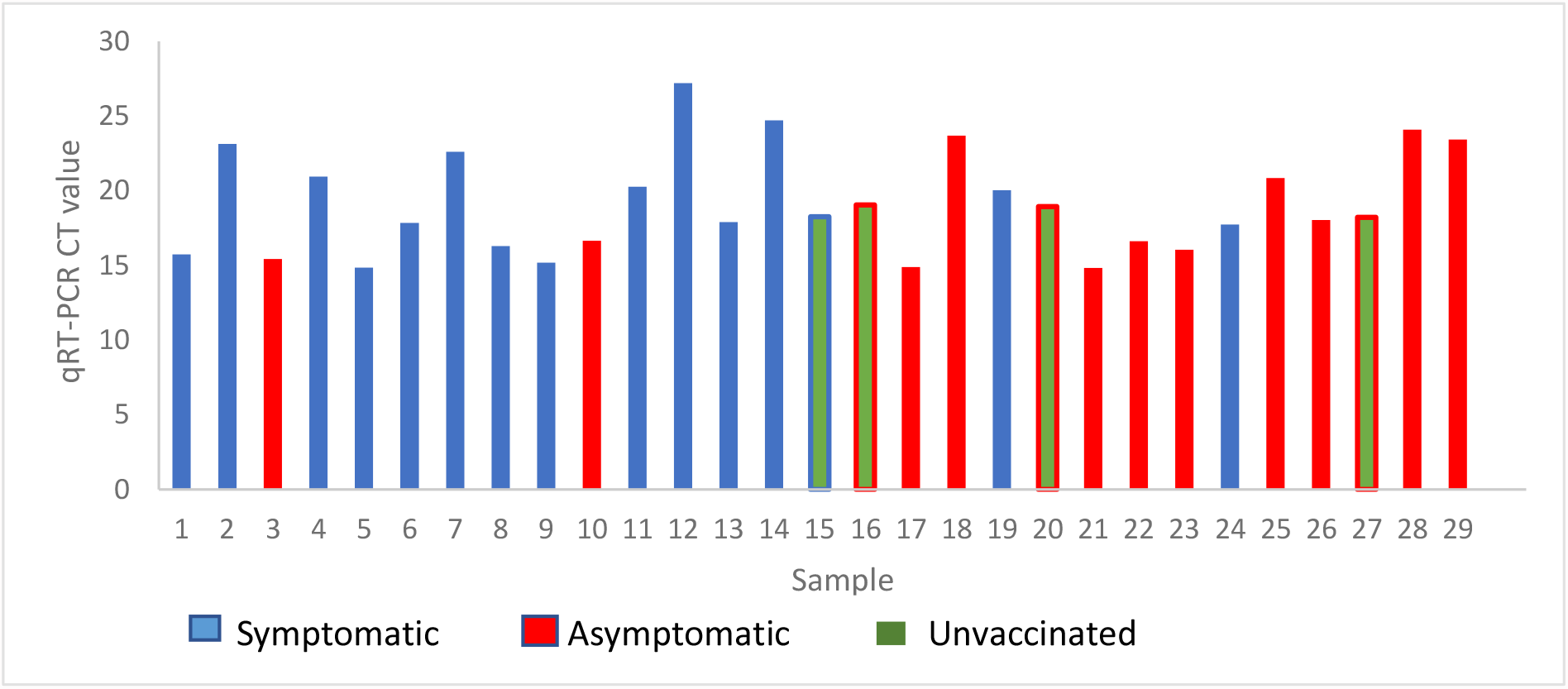
Distribution of Ct values in the clinical samples from vaccinated or unvaccinated infected by Omicron or Delta variant.

**Figure 2.**
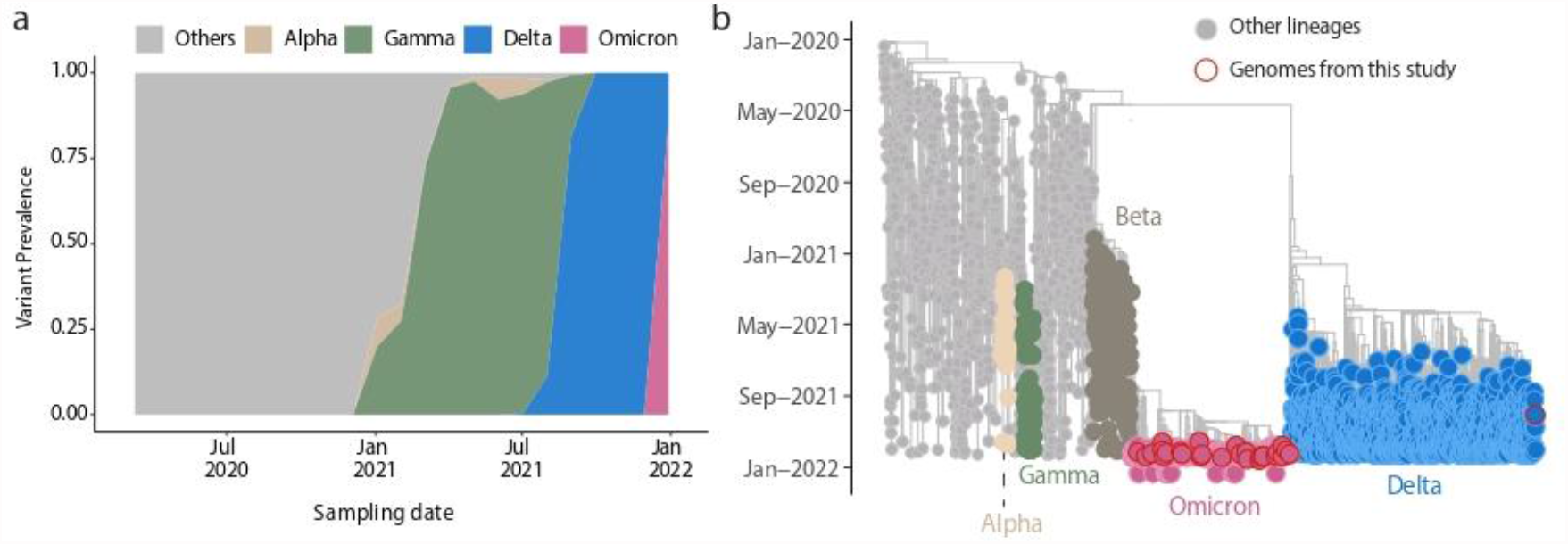
Genomic epidemiology of the SARS-CoV-2. Omicron variant in Salvador, Bahia, Northeast Brazil. A) Dynamics of the SARS-CoV-2 epidemic in Bahia showing the progression in the proportion of circulating variants in the state over time, with the rapid replacement of the Delta by the Omicron variant; B) Time-resolved maximum likelihood phylogenetic tree including the newly (n=29) Omicron isolates obtained in this study plus n = 4,249 representative SARS-CoV-2 genomes collected up to January 16^th^, 2021. Alpha, Beta, Gamma, Delta and Omicron VOCs are highlighted in the tree. Newly Omicron and Delta genomes obtained in this study are highlighted with a red border. Genomes of other lineages are showed in grey.

## Discussion

Since the start of the COVID-19 pandemic in December 2019, efforts have been directed to the development of vaccines against SARS-CoV-2. Given this global situation, WHO has authorized the use of several types of vaccines utilizing mRNA technology, adenoviruses, and inactivated viruses^1^.

Recent reports confirm that vaccines currently in use do not protect against infection, but they do greatly reduce the severity of cases. Indeed, countries that have achieved vaccination coverage in at least 50% of their population managed to decrease the mortality rate. However, the current scenario shows the tendency of vaccinated individuals becoming infected with emerging VOCs that cannot be neutralized by the immunity generated from these vaccines^2^. The new variants have emerged as a result of increased viral circulation, primarily in countries where vaccine coverage is low, which quickly spread worldwide. Examples of these variant leaks include Delta in India and Omicron in South Africa.

Our results reinforce the notion that virus migration generally follows national and international patterns of human mobility, facilitating the spread of emerging VOCs not only within countries, but also globally. The identification of SARS-CoV-2 suspected cases via genome-wide sequencing in the state of Bahia also revealed this, and as of 2020, co-circulation of three different VOCs such as Gamma (P.1), Delta, and Omicron appears to have dominated the epidemiological history in the state. The Alpha variant currently remains at a very low frequency (less than 1%) in the state, consistent with the effectiveness of SARS-CoV-2 vaccines against it while being less effective against new VOCs^5^Initially, it was considered in the literature that reaching herd immunity (≥70%) would prevent the circulation of the virus in the population; this fact is now being questioned. The countries that have reached or surpassed this percentage (England, Israel, and Brazil) have high infection rates since the emergence of Omicron^20,21^. In our state, 74.9 % of the vaccination coverage has currently been reached with two doses, but our work demonstrated breakthrough infection by Omicron with the high viral load even in double or triple vaccinated individuals. Moreover, asymptomatic vaccinated individuals can still spread the virus to other people.

One limitation of our work is not knowing the level of protective immunity of those who are currently vaccinated; however, even with three doses, there have been cases of viral infection. Dose reinforcements in countries such as England and Israel with a four-dose regimen raise doubts as to whether they will solve the situation of viral recrudescence for new VOCs in circulation^22,23^.

It is important to acknowledge the limitations of these vaccines and to urgently develop new vaccines to emergent VOCs, as is the case of the vaccine for Influenza; going through new doses of the same vaccines is “more of the same”.

Measures to control the use of face masks, social distancing, and crowd avoidance continue to be effective, but these need to be combined with new immunogens that prevent viral infection (an elementary concept for the approval of vaccines for widespread use).

## Data Availability

All data produced in the present work are contained in the manuscript

## Acknowledgments

This work was supported by Fundação de Apoio a Extensão e Pesquisa -Bahia-Brazil (FAPESB) GRANT 2020 /COVID 19; MG is supported by Fundação de Amparo à Pesquisa do Estado do Rio de Janeiro (FAPERJ) (E-26/202.248/2018(238504) and by the CRP-ICGEB RESEARCH GRANT 2020 Project CRP/BRA20-03, Contract CRP/20/03.

## Data availability statement

Newly generated SARS-CoV-2 sequence have been deposited in GISAID under Accession Numbers: EPI_ISL_9265729, EPI_ISL_9265745, EPI_ISL_9265746, EPI_ISL_9265743, EPI_ISL_9265744, EPI_ISL_9265749, EPI_ISL_9265728, EPI_ISL_9265747, EPI_ISL_9265748, EPI_ISL_9265730, EPI_ISL_9265752, EPI_ISL_9265731, EPI_ISL_9265753, EPI_ISL_9265750, EPI_ISL_9265751, EPI_ISL_9265734, EPI_ISL_9265756, EPI_ISL_9265735, EPI_ISL_9265732, EPI_ISL_9265754, EPI_ISL_9265733, EPI_ISL_9265755, EPI_ISL_9265738, EPI_ISL_9265739, EPI_ISL_9265736, EPI_ISL_9265737, EPI_ISL_9265741, EPI_ISL_9265742, EPI_ISL_9265740.

